# Clinical characteristics of 34 COVID-19 patients admitted to ICU in Hangzhou, China

**DOI:** 10.1101/2020.04.12.20062604

**Authors:** Yi Zheng, Lijun Sun, Mi Xu, Jian Pan, Yuntao Zhang, Xueling Fang, Qiang Fang, Hongliu Cai

## Abstract

**Introduction:** The purpose of the study was to summarize the clinical and laboratory characteristics of the coronavirus disease 2019 patients admitted to intensive care unit.

**Methods:** We tracked the data until March 5, 2020. The cases in our cohort were divided into cases only received noninvasive ventilation (NIV) and cases required invasive mechanical ventilation (IMV). The characteristics between the two groups were compared.

**Results:** 34 cases were included in the study. The complications rate (including, acute liver injury, acute cardiac injury and acute kidney injury) were higher in IMV cases. Lymphocytopenia and neutrophilia occurred in most cases in both groups on the admission day, however, lymphocyte levels dropped progressively and more severe lymphopenia occurred in IMV group. Increased amounts of plasma IL-6 and IL-10 were found in both groups on the admission day, the progressive decrease of which occurred in NIV cases rather than IMV cases, and the levels were higher in IMV cases during hospitalization.

**Conclusions:** Lymphocytopenia, neutrophilia, and increase of IL-6 and IL-10 occurred in SARS-CoV-2 infected patients in ICU, however, the dynamics of those were significantly different in IMV cases and NIV cases during hospitalization.

## Introduction

In December 2019, a series of acute respiratory illness of unknown cause, now known as Coronavirus Disease 2019 (COVID-19) caused by SARS-Cov-2, emerged in Wuhan, Hubei, China. SARS-Cov-2 was classified in the beta-coronavirus 2b lineage. It has rapidly and widely spread in China and many other countries, causing an outbreak of COVID-19, and has caused great economic loss and mental health stress.

As of 10 March 2020, the cumulative number of confirmed cases in China is more than 80000, of which the death toll has exceeded 3000 [1]. COVID-19 is now the third lethal illness caused by coronavirus following severe acute respiratory syndrome (SARS) [2] and Middle East respiratory syndrome (MERS) [3]. One third of COVID-19 patients were admitted to ICU. As the epidemic progresses, the treatment of severe and critical patients is the key to success of the epidemic battle. The COVID-19 fatality rate between Hubei and non-Hubei are significant differences [1]. The dynamic analysis of the characteristics of COVID-19 patients in ICU of non-Hubei area may have better guiding significance for the future severe patients

We intend to describe clinical and laboratory characteristics, treatment, and outcomes of confirmed COVID-19 patients admitted to ICU in a tertiary teaching hospital in Hangzhou, a place far away from Wuhan in China. The Department of critical care medicine in this hospital is a national key clinical specialty and regional critical diagnosis and treatment center in Zhejiang province. During the COVID-19 epidemic, some severe patients transferred from the surrounding cities were treated in the hospital. The novel coronavirus pneumonia here is characterized by regional representation.

## Methods

### Participants and Study Design

For this retrospective, single-centre study, we analysed patients from Jan 22, 2020, to Mar 5, 2020, who had been diagnosed with SARS-CoV-2 pneumonia, according to the Chinese National Health Commission guidance[4], in ICU at the First Affiliated Hospital, College of Medicine,Zhejiang University, Hangzhou, which is a designated hospital for COVID-19 treatment. Oral consent was obtained from the authorized person. This study was approved by the National Health Commission of China and Ethics Commission of the First Affiliated Hospital, College of Medicine,Zhejiang University (IIT20200077A). Conditions for admission to ICU in our research: dyspnea and respiratory rate ≥ 30 times/min, oxygen saturation ≤ 93% at rest without oxygen inhalation, PaO2/FiO2 (P/F) ≤ 300mmHg (1mmHg=0.133kPa), other organ dysfunction such as shock.

### Data Collection

Two researchers independently reviewed patients’ medical records for epidemiological, demographic, clinical, laboratory, management, and outcome data which were collected until March 5, 2020. The time of disease onset was defined as the day when any related symptom was noticed which were ascertained through researcher communicating with patients or their families. Acute respiratory distress syndrome (ARDS) was identified according to the Berlin definition [5]. Acute kidney injury (AKI) was identified according to the KDIGO Clinical Practice Guidelines [6]. Acute liver injury defined as an increase in alanine aminotransferase (ALT) over two times the upper limit of the normal range (ULN) or conjugated bilirubin or a combined increase in aspartate aminotransferase (AST), alkaline phosphatase and total bilirubin provided that one of them was above two times ULN [7]. Cardiac injury followed the definition described in the previous study [8]. Noninvasive ventilation included nasal oxygen therapy, mask oxygen inhalation and HFNC. The durations from onset of disease to first positive nucleic acid testing of respiratory tract specimen, ICU admission, ARDS, HFNC, IPPV, ECMO were recorded.

### Laboratory Confirmation and Treatment

Laboratory tests, including complete blood count, serum biochemistry, coagulation profile, lactate dehydrogenase, C-reactive protein etc. were conducted in the hospital laboratory. Plasma cytokines from all patients (IL-2, IL-4, IL-6, IL-10, TNF-α, IFN-γ) were detected by ELISA method according to the manufacturer’s instructions. T, NK and B lymphocyte cell counts were tested by flow cytometry. Respiratory specimens of all patients, including sputum, pharyngeal swabs, bronchoalveolar lavage fluid, or bronchial aspirates were tested for SARS-Cov-2, using real time reverse transcription polymerase chain reaction assays [8]. Antiviral treatment were used in most patients. The majority of patients received treatment with corticosteroid and gamma globulin, the specific dose of which was determined by the chief physician according to the patient’s condition. Antibiotics were empirically administered.

Indications for HFNC: ratio of PaO2 to FiO2 (P/F) < 200, resting respiratory rate was more than 20 per minute and the patient was conscious and cooperative. The timing of invasive ventilation: P/F < 150, resting respiratory rate was more than 30 per minute, uncooperative patients, serious complications such as shock. Indications for ECMO [9]: severe hypoxemia (e.g., P/F < 80, despite the application of high levels of positive end expiration pressure [typically 15–20 cm of water]) for at least 6 hr in patients with potentially reversible respiratory failure, uncompensated hypercapnia with acidemia (pH < 7.15) or excessively high end-inspiratory plateau pressure (>35–45 cm of water, according to the patient’s body size) despite the best accepted standard of care for management with a ventilator.

### Statistical Analysis

Continuous variables were described medians with interquartile ranges and compared with the Mann-Whitney U test. Categorical variables were described using frequency rates and percentages and compared by χ^2^ test or Fisher’s exact test. A two-sided α of less than 0.05 was considered statistically significant. All analyses were performed using SPSS software, version 22.0. The patients in our cohort were divided into cases only received NIV and cases required IMV, because the way of respiratory support was independently associated with the process and outcome [10, 11]. The clinical and laboratory characteristics between the two groups were compared

## Results

By 5 March 2020, 35 cases infected with SARS-CoV-2 in ICU at the First Affiliated Hospital, College of Medicine,Zhejiang University, Hangzhou were investigated. Of the 35 cases, an infected pregnant woman admitted to ICU after caesarean section was excluded because she was an exception and didn’t measure up to the ICU admissions standard. Finally, 34 cases were included in the study. None was residents of Wuhan City. No patients had direct exposure to Huanan seafood market and only 9 patients had been exposed to individuals with confirmed SARS-Cov-2 infection or clustering onset. 17 cases had history of sojourn in epidemic area or close contacted with people from the epidemic area. 8 had no history of any of the above. The median duration from onset of symptoms to the time of first nucleic acid test positive was 4 (IQR 1-7) days. The median duration from onset of symptoms to ICU admission was 10.0 (7·0–11·3) days, to HFNC was 10 (7-11) days in 27 patients, to IMV was 11(8-12) days in 15 cases, to ECMO was 23(18-29) days in 11 patients. (Fig 1)

**Fig 1.**
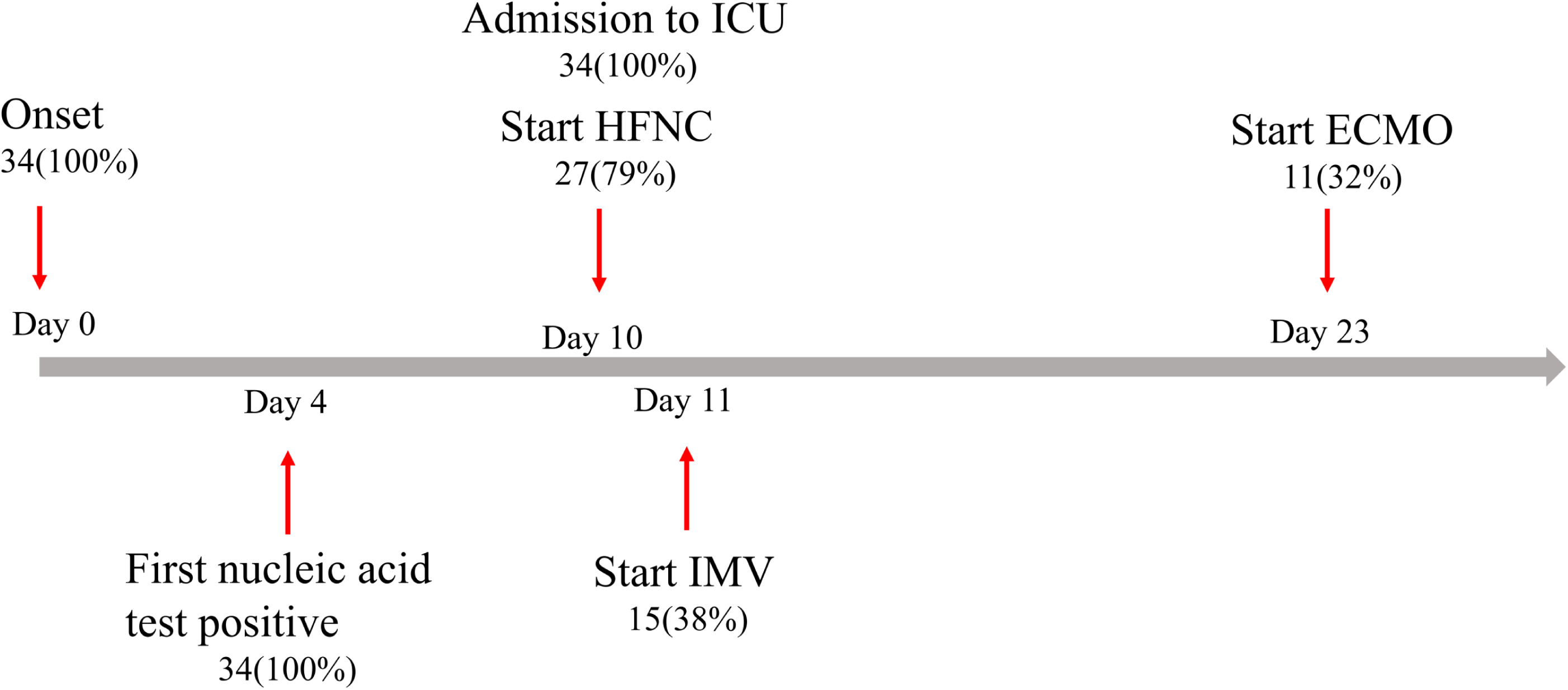
Timeline of COVID-19 cases after onset of illness. ICU: Intensive care unit, HFNC: High-flow nasal cannula, IMV: invasive mechanical ventilation, ECMO: extracorporeal membrane oxygenation

The median age was 66 years (IQR 58-76 years) and 23 (67.6%) were men. 24 (70.6%) patients had chronic diseases, including hypertension (64.7%), diabetes (23.5%), cardiovascular disease (11.8%), chronic obstructive pulmonary disease (5.9%), chronic liver disease (11.8%), chronic kidney disease (5.9%). The most common symptoms at onset of illness were fever (58.8%), dry cough (20.6%), and expectoration (29.4%; table 1), relative less common initial symptoms were myalgia (14.7%), fatigue (5.9%), diarrhea (5.9%), headache (5.9%). The proportion of different comorbidities and the distribution of first symptoms did not differ obviously. (Table 1)

**Table 1:** Demographics and baseline characteristics of patients with covid-19.

| **Characteristics** | **All cases (1)**<br>**(n=34)** | **IMV (2)**<br>**(n=15)** | **NIV (3)**<br>**(n=19)** | **P value**<br>**(2) vs (3)** |
| --- | --- | --- | --- | --- |
| Age, years | 66[58,76] | 71[60,83] | 66[51,72] | 0.176 |
| sex |  |  |  |  |
| Male | 23(67.6%) | 11(73.3%) | 12 (63.2%) | 0.715 |
| Female | 11(32.4%) | 4(26.7%) | 7(36.8%) |  |
| Comorbidities |  |  |  |  |
| Any | 24(70.6%) | 11(73.3%) | 13(68.4%) | 1.000 |
| Hypertension | 22(64.7%) | 10(66.7%) | 12(63.2%) | 1.000 |
| Diabetes | 8(23.5%) | 4(26.7%) | 4(21.1%) | 1.000 |
| Cardiovascular disease | 4(11.8%) | 3(20%) | 1(5.3%) | 0.299 |
| COPD | 2(5.9%) | 1(6.7%) | 1(5.3%) | 1.000 |
| Chronic liver disease | 4(11.8%) | 1(6.7%) | 3(15.8%) | 0.613 |
| Chronic kidney disease | 2(5.9%) | 1(6.7%) | 1(5.3%) | 1.000 |
| Signs and symptoms |  |  |  |  |
| Fever | 20(58.8%) | 11(73.3%) | 9(47.4%) | 0.171 |
| Dry cough | 7(20.6%) | 2(13.3%) | 5(26.3%) | 0.426 |
| Expectoration | 10(29.4%) | 7(46.7%) | 3(15.8%) | 0.068 |
| Fatigue | 2(5.9%) | 1(6.7%) | 1(5.3%) | 1.000 |
| Diarrhea | 2(5.9%) | 0(0%) | 2(10.5%) | 0.492 |
| Myalgia | 5(14.7%) | 2(13.3%) | 3(15.8%) | 1.000 |
| Headache | 2(5.9%) | 2(13.3%) | 0(0%) | 0.187 |
| Vital signs at ICU admission |  |  |  |  |
| Temperature (°C) | 37.3[37,38] | 37.4[37,38.1] | 37.2[37,38] | 0.889 |
| Respiratory rate | 23[19,29] | 25[22,32] | 20[16,26] | 0.060 |
| Heart rate | 79[67,88] | 67[62,91] | 80[70,87] | 0.509 |
| MAP (mm Hg) | 89[84,99] | 85[81,94] | 94[85,100] | 0.022 |

Data are median [IQR], n (%), p values comparing NIV cases and IMV cases are from Mann-Whitney U test or Fisher's exact test. ICU: Intensive care unit, NIV: noninvasive ventilation, IMV: invasive mechanical ventilation, IQR: interquartile range, COPD: chronic obstructive pulmonary disease

The blood cells counts on admission showed that leukocytosis (white blood cell count more than 10 × 10^9^/L, 15 [44.1%] cases) occurred in nearly half of the patients, while, the incidence of leucopenia (white blood cell count less than 4 × 10^9^/L, 2 [5.9%] cases) is very low. Lymphopenia (lymphocyte count < 0.8 × 10^9^/L, 22 [66.4%] patients) occurred in more than half of the patients on admission. Platelet count in NIV group was higher than in IMV group, which were in normal range in most patients of both groups. No significant differences were found between NIV and IMV groups in terms of biochemical indexes, inflammation or immune indicators and coagulation indices on admission to hospital. (Table 2)

**Table 2.** Laboratory findings in patients with covid-19 on admission to hospital.

| Variables | Normal Range | All patients (1)<br>(n=34) | IMV (2)<br>(n=15) | NIV (3)<br>(n=19) | P value<br>(2) vs (3) |
| --- | --- | --- | --- | --- | --- |
| WBC count ($\times 10^9/L$) | 4.0-10.0 | 8.9[5.3,14.3] | 6.2[4.4,13.5] | 9.3[6.6,16] | 0.160 |
| < 4 |  | 2(5.9%) | 1(6.7%) | 1(5.3%) |  |
| 4 - 10 |  | 17(50%) | 8(53.3%) | 9(47.7%) | 0.674 |
| > 10 |  | 15(44.1%) | 6(40%) | 9(47.7%) |  |
| Neutrophil count ($\times 10^9/L$) | 2.0-7.0 | 7.8[4.3,13.2] | 5.7[3.6,12.8] | 8[5.5,14.8] | 0.223 |
| Lymphocyte count ($\times 10^9/L$) | 0.8-4.0 | 0.55[0.48,0.9] | 0.5[0.4,0.8] | 0.6[0.5,0.9] | 0.539 |
| < 0.8 |  | 22(64.4%) | 10(66.7%) | 12(63.2%) |  |
| $\geq 0.8$ | | 12(35.3%) | 5(33.3%) | 7(36.8%) | 1.000 |
| Platelet count ($\times 10^9/L$) | 101-320 | 178[147,196] | 156[111,176] | 191[172,201] | 0.012 |
| ALT U/L | 7-40 | 20[20,30] | 21[14,31] | 19[15,27] | 0.768 |
| Total bilirubin, $\mu\text{mol/L}$ | 0.0-21.0 | 12[7.6,18.6] | 11.8[7.3,18.5] | 12.1[8,18.7] | 0.876 |
| BUN, mmol/L | 3.10-8.80 | 7[5.4,10.5] | 7.5[6,14.2] | 6.6[5.0,8.1] | 0.111 |
| Creatinine, $\mu\text{mol/L}$ | 41-81 | 84[67,104] | 84[67,106] | 84[67,99] | 0.445 |
| C-reactive protein, mg/L | 0.00-8.00 | 48[27,88] | 50[39,52] | 46[27,103] | 0.986 |
| D-dimer, mg/L | 0-700 | 606[383,1109] | 675[433,1613] | 467[370,1073] | 0.405 |
| Interleukin- 6, pg/mL | 0.00-6.61 | 47[24,81] | 65[38,202] | 38[12,72] | 0.054 |
| Interleukin -10, pg/mL | 0.00-2.31 | 6.7[4.4,9] | 7[5.2,9.7] | 5.5[3.1,8.8] | 0.258 |

Data are median [IQR], n (%), p values comparing NIV cases and IMV cases are from Mann-Whitney U test or Fisher's exact test. NIV: noninvasive ventilation, IMV: invasive mechanical ventilation, IQR: interquartile range, WBC: white blood cell count, ALT: alanine aminotransferase, AST: aspartate aminotransferase BUN: blood urea nitrogen, LDH: lactate dehydrogenase

Only one patient did not receive antiviral and systematic corticosteroid treatment. 30 (88.2%) were given empirical antibiotic treatment, and 27 (79.4%) were given gamma globulin treatment. 18 patients (52.9%) received HFNC without escalation of respiratory support. 15 (44.1%) received IMV and ECMO was required in 11 patients (32.4%). Continuous renal replacement therapy (CRRT) was conducted in 5(14.7%) cases. Common complications among the 34 ICU patients included ARDS (33 [97.1%]), acute liver injury (13 [38%]), acute cardiac injury (13 [38%]), and AKI (7[20.6%]). The complications rates (including, acute liver injury, acute cardiac injury and AKI) were significantly higher and discharge rate was notably lower in IMV cases, compared with NIV patients. (Table 3)

**Table 3.** Treatments, complications and outcomes in patients with covid-19 during hospitalization.

|  | **All cases (1)**<br>**(n=34)** | **IMV (2)**<br>**(n=15)** | **NIV (3)**<br>**(n=19)** | **P value**<br>**(2) vs (3)** |
| --- | --- | --- | --- | --- |
| **Treatments** |  |  |  |  |
| Antiviral therapy | 33(97.1%) | 15(100%) | 18(94.7%) | 0.559 |
| Glucocorticoid therapy | 33(97.1%) | 15(100%) | 18(94.7%) | 0.559 |
| Antibiotics | 30(88.2%) | 15(100%) | 15(78.9%) | 0.084 |
| Gamma globulin | 27(79.4%) | 14(93.3%) | 13(68.4%) | 0.085 |
| HFNC | 18(52.9%) | 0(0%) | 18(94.7%) | < 0.0001 |
| IMV | 15(44.1%) | 15(100%) | 0(0%) | < 0.0001 |
| ECMO | 11(32.4%) | 11(73.3%) | 0(0%) | < 0.0001 |
| CRRT | 5(14.7%) | 4(26.7%) | 1(5.3%) | 0.104 |
| **Complications** |  |  |  |  |
| ARDS | 33(97.1%) | 15(100%) | 18(94.7%) | 0.559 |
| AKI | 7(20.6%) | 6(40%) | 1(5.3%) | 0.019 |
| Acute liver injury | 14(41.2%) | 9(60%) | 5(26.3%) | 0.051 |
| Acute cardiac injury | 13(38%) | 10 (66.7%) | 3(15.8%) | 0.003 |
| **Outcomes** |  |  |  |  |
| Discharge | 20(58.8%) | 2(13.3%) | 18(94.7%) | < 0.0001 |
| Death | 0 | 0 | 0 | 1.000 |

Data are n (%), p values comparing NIV cases and IMV cases are from Fisher's exact test. NIV: noninvasive ventilation, IMV: invasive mechanical ventilation, HFNC: High-flow nasal cannula, ECMO: extracorporeal membrane oxygenation, CRRT: continuous renal replacement therapy, ARDS: acute respiratory distress syndrome, AKI: acute kidney injury

Dynamic changes of the main laboratory indicators during COVID-19 progression including blood cells counts and biochemical parameters, coagulation profile and inflammatory cytokines were followed from day 1 to day 9 or discharged day after admission at 1-day interval. White blood cell counts and neutrophil counts were at high levels during hospitalization, and showed declining trends on discharge day in NIV patients. During hospitalization, most patients had marked lymphopenia, and IMV patients developed more severe lymphopenia over time, but lymphocytes returned to normal levels in NIV cases on discharge day. Flow cytometry showed that T lymphocytes counts were further away from the lower normal limit, compared to B lymphocytes in inpatients. T lymphocytes counts stayed low during hospitalization in IMV patients, however, the counts gradually rose to normal level until discharge in a part of NIV patients. Natural killer cells counts in most inpatients were far from the lower normal limit, and the counts progressive declined in IMV cases rather in NIV cases during hospitalization. Platelet counts and hemoglobin levels were higher in NIV cases than those in IMV cases, and hemoglobin levels dropped progressively in IMV cases during hospitalization. C-reactive protein levels in both groups showed gradual downward trend during hospitalization, however, that in NIV cases descended to a lower level at a faster rate. ALT and D-dimer levels showed upward trends in both groups during hospitalization, while, for total bilirubin and blood urea nitrogen, the upward trends only occurs in IMV cases. The progressive decrease in LDH, IL-6 and IL-10 levels occurred in NIV cases rather than IMV cases during hospitalization, and the levels were higher in IMV cases than in NIV cases. (Fig 2 and Fig 3)

**Fig 2.**
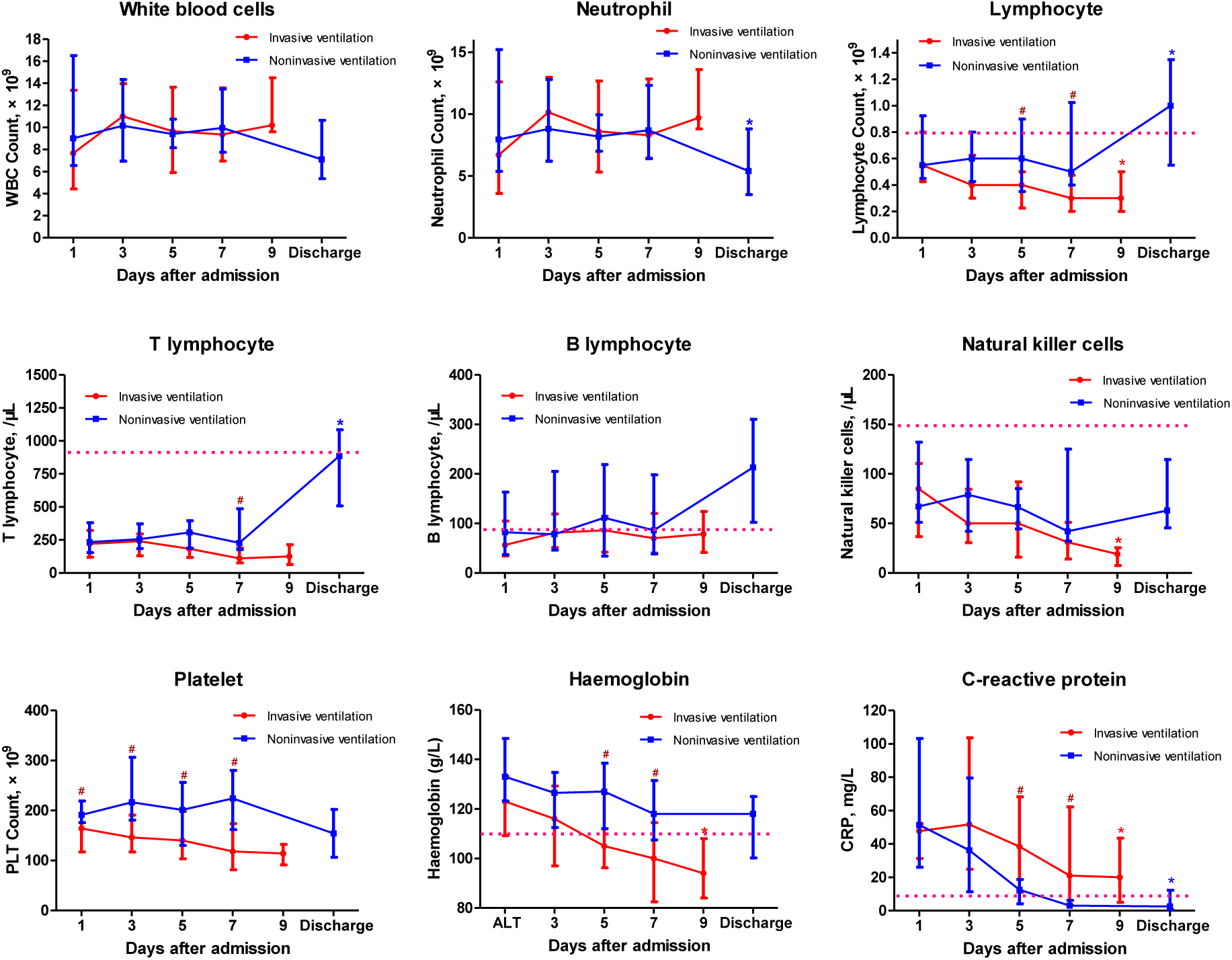
Timeline charts illustrate the laboratory parameters in 34 patients with COVID-19(19 NIV cases and 15 IMV cases) every other day based on the days after admission. The dotted line shows the lower normal limit in lymphocyte, T lymphocyte, B lymphocyte, natural killer cell and haemoglobin and shows the upper normal limit in CRP. ^*^p < 0.05 vs. day 1 within group. ^#^p < 0.05 between two groups on the same day.

**Fig 3.**
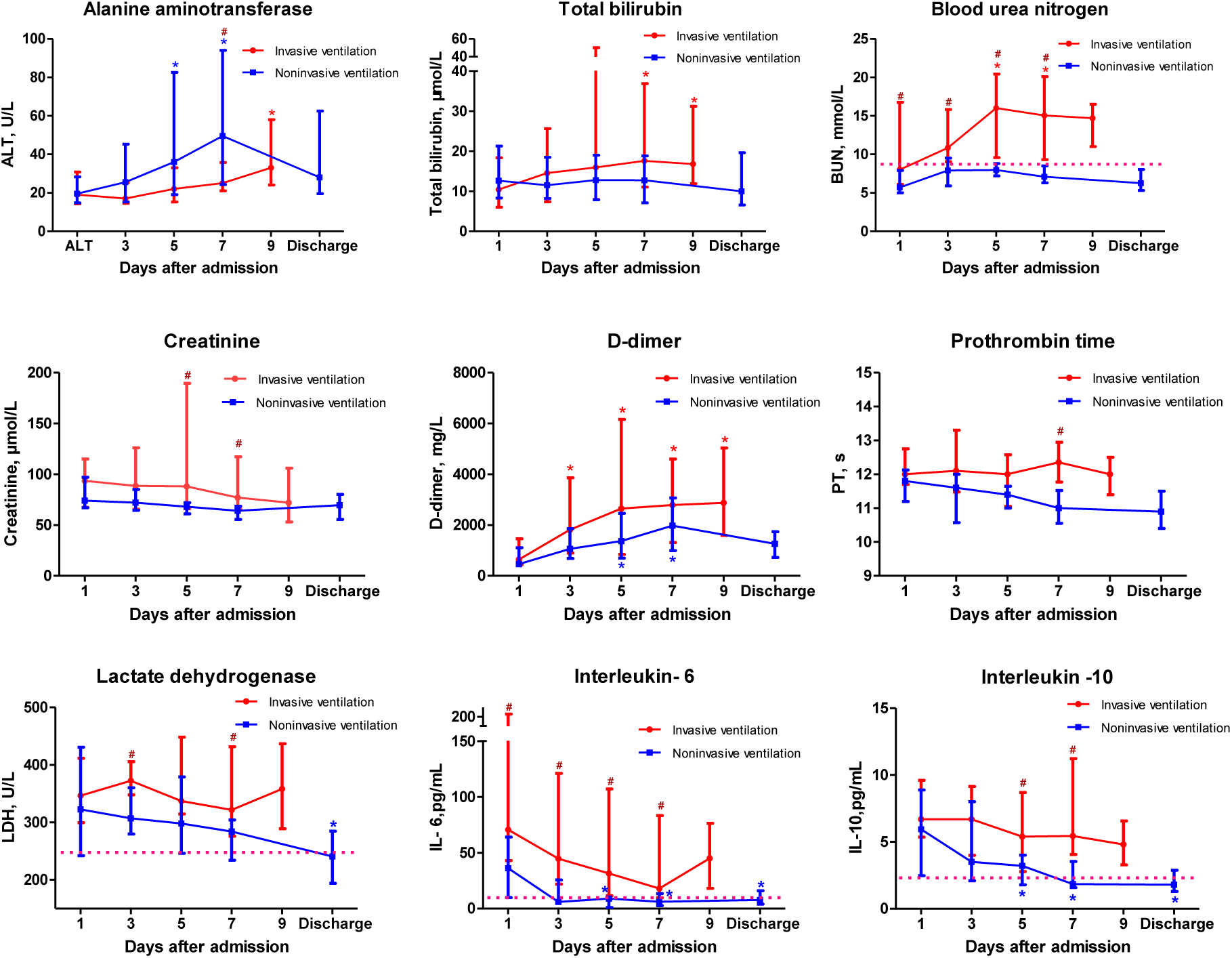
Timeline charts illustrate the laboratory parameters in 34 patients with COVID-19(19 NIV cases and 15 IMV cases) every other day based on the days after admission. The dotted line shows the upper normal limit in BUN, LDH, IL-6, IL-10. ^*^p < 0.05 vs. day 1 within group. ^#^p < 0.05 between two groups on the same day.

## Discussion

We report a cohort of 34 ICU patients with laboratory-confirmed SARS-CoV-2 infection, characterised by severe or critical coronavirus pneumonia. 67.6% patients were men, with a median age of 66 years (IQR 58-76) and the oldest patient, a 94 years old man has been discharged. Of the 34 patients, 19 patients (55.9%) only received noninvasive respiratory support (including nasal oxygen therapy in one case and HFNC in 18 patients) and 15 (44.1%) required IMV. By the end of Mar 5 2020, the complications rate (including, acute liver injury, acute cardiac injury and AKI) and discharge rate were significantly different between the two groups.

More than 100 COVID-19 patients were admitted to our designated hospital. Patients in general isolation ward were mild to moderate, and most ICU patients without IMV required have got better and discharged. However, the majority of ICU cases with IMV required are still under respiratory support, including 11 ECMO operating. So, the main battlefield of the epidemic war is in ICU, and the key to the success of the epidemic fighting is striving to improve and perfect the management and treatment of severe and critical patients. Most common complication was ARDS (33 [97.1%] of 34 patients) in the research, so, respiratory support is essential. By now, no antiviral treatment has been proven to be effective for coronavirus infection. 97.1% infected patients received single or combined antiviral, including baloxavir, favipiravir, darunavir, arbidol, or interferon alpha inhalation. Corticosteroid and gamma globulin were administrated to a part of patient, but the efficacy needs to be further confirmed.

Most patients had neutrophilia in our cohort, which was consistent with the results of Raymond et al in SARS-CoV infected patients [12]. As mentioned in previous studies for critically ill patients with SARS-CoV [13], SARS-CoV-2 [14] and MERS [15, 16] infection, lymphocytopenia is a prominent feature because targeted invasion by viral particles causes lymphocyte destruction. Lymphocytopenia occurred in more than 60% patients in our cohort on the first day of admission. As the disease progresses, more patients developed lymphocytopenia. In the IMV group with more severe illness, lymphocyte levels dropped progressively and more severe lymphopenia occurred compared to no-IMV group, which suggested that the severity of lymphocytopenia reflects the severity of SARS-CoV-2 infection. As the research of SARS-CoV infected cases shows that patients with more severe clinical illness, or patients who died, had significantly more profound lymphopenia [17]. The lymphocyte counts can gradually rise to normal level until discharge in a part of patients without IMV in our cohort. Lymphopenia was prolonged, and the lymphocyte returned towards normal after five weeks of illness in patients with SARS-CoV [17] infection or within 2–3 weeks after the disease onset in cases with severe pandemic H1N1 influenza A [18]. Further analysis in our cohort showed that T lymphocytopenia was more pronounced than B lymphocytopenia which was similar to previous study in Patients with SARS [19]. The mechanism behind the dynamics is not clear, which need to be confirmed in cell and animal experiments.

We noted that SARS-CoV-2 infection caused increase in plasma IL-6 levels, which were consistently at a high levels in IMV cases in our cohort. Early studies also have shown that increased concentrations of proinflammatory cytokines (eg, IL-6, TNFα and IFNγ) in SARS patients [20] and MERS-CoV cases [21]. Increased amounts of anti-inflammatory cytokine, IL-10, in plasma was found in SARS-CoV-2 infected patients with IMV, which differs from SARS-CoV infection [12], but supported by Huang et al. [8] in patients with the same disease. BUN is a key element reflecting the intricate interrelation between nutritional status, protein metabolism and renal situation of the patient [22]. BUN were significantly elevated in IMV patients compared with NIV cases which may cause by high catabolism. The characteristics of inflammation and metabolism, combined with lymphocytopenia, suggested that we have to be on the alert of the existence of persistent inflammation-immunosuppression and catabolism syndrome (PICS) in IMV cases. PICS, often leading to secondary infections and/or viral reactivation in the critically ill, has been associated with increased morbidity and mortality [23-25].

This study has several limitations. Firstly, limited samples in our cohort are analyzed. A previous report enrolled 52 critically ill patients with SARS-CoV-2 infection from Wuhan Jin Yin-tan hospital. However, in places outside Hubei province, the total number of cases and the number of critical patients are relatively small. Secondly, most IMV patients had not been discharged at the time of manuscript submission, so we can’t use survival or death as a clinical end point. The case mortality rate and other outcome indicators need to be reported later.

## Conclusion

In this single-center case series of 34 ICU patients with SARS-CoV-2 infected in Hangzhou, China, 97.1% of patients were complicated by ARDS, 44.1% received IMV, 55.9% only needed noninvasive respiratory support. Compared with cases in NIV group, patients received IMV were easier to complicate with organs injury, developed more severe lymphopenia, and had higher inflammatory markers.

## Data Availability

The data will be made available to others on reasonable
requests to the corresponding author.

## List of abbreviations

COVID-19: coronavirus disease 2019
ICU: Intensive care unit
NIV: noninvasive ventilation
IMV: invasive mechanical ventilation
IQR: interquartile range
COPD: chronic obstructive pulmonary disease
HFNC: High-flow nasal cannula
ECMO: extracorporeal membrane oxygenation
LDH: lactate dehydrogenase
SARS: severe acute respiratory syndrome
MERS: Middle East respiratory syndrome
P/F: PaO2/FiO2
ARDS: acute respiratory distress syndrome
AKI: acute kidney injury
ALT: alanine aminotransferase
ULN: upper limit of the normal range
AST: aspartate aminotransferase
IPPV: Invasive positive pressure ventilation
CRRT: continuous renal replacement therapy
BUN: blood urea nitrogen
PICS: persistent inflammation-immunosuppression and catabolism syndrome

## Declarations

### Acknowledgements

Not applicable.

### Authors’ contributions

HLC and YZ conceived and designed the study, and take responsibility for the integrity of the data and the accuracy of the data analysis. LJS, MX and JP collected the data. YZ, XLF and YTZ did the analysis. HLC and YZ drafted the paper. All authors agree to be accountable for all aspects of the work in ensuring and have read and approved the manuscript.

### Availability of data and materials

Not applicable

### Ethics approval and consent to participate

The study was conducted in accordance with the principles of the Declaration of Helsinki, and Institutional Review Board approval has been obtained.

### Consent for publication

Not applicable

### Author details

Not applicable

